# Sexual health empowerment interventions for migrant women: a systematic review of the literature

**DOI:** 10.1101/2025.02.21.25322665

**Authors:** Lisa Ouanhnon, Laure Daumail, Marie Kerihuel, Aurore Palmaro, Julie Dupouy, Marie-Anne Durand

## Abstract

**Introduction:** Migrant women are suffering from inequalities in access to care and prevention, quality of care, and in particular, sexual health care. As part of an overall project to develop a primary care intervention aimed at empowering migrant women in sexual health, the first objective will be to review the characteristics of sexual health empowerment interventions targeting migrant women. The second objective will be to assess the effectiveness of these interventions.

**Methods and analysis:** A systematic review will be conducted across PubMed, Cochrane, Embase, Psychinfo/ Psycharticles, Web of science, Cairn, as well as grey literature, from inception until 01/01/2023. We will include randomized clinical trials, quasi-experimental, and qualitative studies published in any language before 01/01/2023, assessing the effect of a sexual health empowerment intervention targeted at migrant women and meeting the selection criteria. Screening of articles and data extraction will be conducted by two independent researchers. Disagreements will be resolved by a third reviewer. The quality of all included studies will be assessed using validated tools according to the study design. Results will be reported using the Preferred Reporting Items for Systematic Reviews and Meta-Analysis guidlines, and a narrative synthesis will be conducted.

**Ethics and diffusion:** This systematic review will involve only secondary data, so ethical approval was not required for this study. The results will be disseminated in peer-reviewed journals and conferences.

## INTRODUCTION

There are currently more than 1 billion people on the move in the world, including 281 million international migrants [1]. An estimated 48% of these migrants are women [2]. The number of people on the move in the world is expected to increase in the coming years, as a result of war, violence and persecution, as well as climate change. For several years now, the health of these populations has been recognized as a social and political priority [3]. Worldwide, the health status of vulnerable migrants is poorer than that of people from host countries [4]. This difference is not intrinsic, but relates to a difference in multiple health determinants (education, income, housing). This difference is also influenced by health conditions acquired during the migration process, and often related to poorly adaptable care systems in host countries.

These inequalities are also found in the field of sexual health. According to the WHO, sexual health is “a state of physical, emotional, mental and social well-being in relation to sexuality; it is not merely the absence of disease, dysfunction or infirmity. Sexual health requires a positive and respectful approach to sexuality and sexual relationships, as well as the possibility of having pleasurable and safe sexual experiences, free of coercion, discrimination and violence.”[5]. Sexual health is a complex, intimate topic, which is particularly difficult to address in the context of language and cultural barriers [6]. Among displaced women, the violence experienced in their country of origin and during their migration also has an impact on their state of health [7]. As a result of these adversities, combined with difficult access to healthcare in host countries, displaced women are exposed to health risks such as delayed diagnosis of cancer due to limited access to screening [8,9], higher rates of sexually transmitted infections (STIs) [10,11], poorer birth control [12,13] and higher rates of perinatal complications [14–17].

To improve the health of vulnerable populations, empowerment interventions are increasingly being developed worldwide [18]. Empowerment is defined by the WHO as “the process by which people gain control over the factors and decisions that shape their lives”.

In sexual health, empowerment refers to the ability to make the decisions necessary to exercise control over one’s sexual life [19,20]. Several interventions, have been developped in a wide variety of social and geographical contexts, concerning STI prevention, organised cancer screening or access to contraception [19,21–23].

In order to reduce existing health inequalities between migrant and French women, we are currently conducting the ‘ENVOL’ research project (Empowerment iN sexual health: an interVention for migrant wOmen in generaL practice), which ultimately aims to develop, evaluate and implement a primary care empowerment intervention in sexual health for migrant women in France. The review of existing interventions designed to empower migrant women worldwide about their sexual health is an essential prerequisite for the subsequent development, evaluation and implementation of this intervention.

## OBJECTIVES

The first objective will be to review the characteristics of sexual health empowerment interventions targeting migrant women. The second objective will be to assess the effectiveness of these interventions.

## METHODS AND ANALYSIS

### Protocol and registration

A systematic review of the literature will be conducted, following Preferred Reporting Items for Systematic Reviews and Meta-Analyses (PRISMA) guidelines [24]. This review protocol has been registered in PROSPERO (number CRD4202440894), and was designed in accordance with the Preferred Reporting Items for Systematic Reviews and Meta-Analyses Protocols (PRISMA-P) checklist.

### Study selection criteria

Inclusion criteria are presented according the PICOS framework: Population, intervention, comparator, outcome, and study design criteria, as described by Satos et al. [25].

#### Type of participants

The review will focus on migrant women aged 15 and over, with no restriction on nationality. Studies focused on population with a particular pathology, a population exclusively or predominantly (i.e. >50%) male, or exclusively or predominantly (i.e. >50%) under 15 years will not be included.

#### Type of intervention

Interventions aimed at empowerment in sexual health will be taken into consideration. These interventions may concern :

- Prevention, screening and treatment of sexually transmitted infections
- Screening for cervical and breast cancer
- Sex education
- Screening and care for sexual violence
- Contraception and birth control

This may include studies with several interventions or with multi-component interventions. Only interventions that have been carried out in upper-middle to high income countries with a healthcare system comparable to the French one.

#### Type of comparator

No restrictions on the type of comparator will be applied.

#### Type of outcome measure

All outcomes concerning the effectiveness of interventions will be considered. Outcomes concerning the acceptability and the feasibility of interventions will be taken into consideration. Studies with economic criteria as the primary outcome will not be included.

#### Type of studies

Randomised controlled trials, pilot randomised controlled trials, quasi-experimental, and qualitative studies will be considered. Any exclusively descriptive study, observational studies, literature reviews, editorials, commentaries, or books will not be included.

### Search strategy

#### Electronic databases

The literature search was performed in PubMed, Cochrane, Embase, Psychinfo/Psycharticles, Web of science and Cairn using search equations, for publication in any language published from inception until 01/01/2023. Additional file indicates the databases and the retrieval formula used for each one.

#### Additional search methods

Two independent researchers will manually search the reference lists of all selected articles and of the systematic reviews on related topics. Two independent researchers will manually search relevant sources of grey literature : ClinicalTrials.gouv, SUDOC, French National Authority for Health; WHO; Institut Convergence Migration; and French College of Gynaecology and Obstetrics.

#### Study selection

Two researchers will independently assess the title and abstract of retrieved records. After agreeing on the list of included articles, they will independently assess the full text articles meeting the inclusion criteria. Disagreements will be resolved by a third researcher. We will review and consider all search results for inclusion using Rayyan, a web application designed for screening systematic review records. Since Rayyan is an online application, data are continuously backed up and managed by the website.

#### Data extraction

The data will be extracted independently by two researchers, using a pre-established, piloted grid, then pooled secondarily. The grid was elaborated using TIDieR (Template for Intervention Description and Replication) criteria.Three researchers will pilot the data extraction grid, using a set of five studies selected for this pilot phase.The final grid will include the author, country, year of publication, type of study and design, study population and sample size, type of intervention (based on the TIDieR checklist, with an additional item on cultural tailoring), comparator (if present), follow-up (if appropriate) and primary and secondary outcomes.

### Quality assessment

The methodological quality of each included study will be assessed independently by the two researchers. For randomized controlled trials, the Cochrane Risk of Bias Tool 2 will be used [26]. For qualitative studies, we will apply the Cheklist Critical Appraisal Skills Program (CASP) [27]. To assess the quality of quasi-experimental studies, the Newcastle-Ottawa quality assessment scale will be used [28].

### Data Analysis

The clinical heterogeneity of the studies included, in terms of types, content of the intervention, target populations, sexual health topic, and outcomes is not expected to allow a quantitative analysis.

Data will be analysed using a narrative synthesis. This synthesis will be inspired by the ‘Guidance on the Conduct of Narrative Synthesis in Systematic Reviews’[29]. A thematic synthesis of the form and content of the interventions will be proposed, as well as their effectiveness. An analytical reading will be carried out to highlight the use of empowerment techniques in the proposed interventions and the inclusion of the concept of empowerment in the outcomes used.

## Data Availability

The data supporting the conclusions of this review will be available upon reasonable request from the corresponding author and with the permission of the sponsor.

## SUPPLEMENTARY INFORMATION

Additional file 1. Search strategy.

## ETHICS AND DIFFUSION

## Ethics

This systematic review will involve only secondary data, so ethical approval was not required. The findings of this systematic review will be submitted for publication in peer-reviewed journals and presented at scientific conferences. This protocol has been registered in PROSPERO (CRD42024408949). The systematic review will be reported according to PRISMA guidelines. All protocol amendments will be reported in the final scientific manuscript of this systematic review.

## Acknowledgements

We would like to thank the bibliographic information specialist Tony Chatelain for his help in selecting the databases and developing the search strategy.

## Contributors

LO, AP, JD and MAD contributed to the conception and design of the study.

LO and AP registered the protocol in the PROSPERO database. LO, AP, MK and LD drafted the protocol.

All authors revised the protocol critically for important intellectual content.

LO, MK, LD, MAD and JD designed the search strategy, and participated in the design of data acquisition, analysis and interpretation.

All authors read and approved the final protocol. LO is the guarantor of this protocol.

## Funding

The sponsor of this research is the university multi-professional health center of Pins-Justaret. This study was supported by a grant from the French Ministry of Health (ReSPir 2021 / N° RESPIR-21-035)

## Competing interests

Marie-Anne Durand has contributed to the development of Option Grid patient decision aids. EBSCO Information Services sells subscription access to Option Grid patient decision aids. She receives consulting income from EBSCO Health, and royalties. No other competing interests declared.

## Patient consent for publication

Not required.

## Patient and public involvement

This review protocol will not directly involve the patients or general public. However, patients will be involved in the later stage of the ongoing project in which this review is included.

## ABBREVIATIONS

CASP: Cheklist Critical Appraisal Skills Program
GP: general practitioner
HIV: Human Immunodeficiency Virus
PRISMA: Preferred Reporting Items for Systematic Reviews and Meta-Analyses
RCTs: Randomized controlled trials
STIs: Sexually Transmitted Infections
TIDieR: Template for Intervention Description and Replication

---

**Table 1.** PICOS criteria for inclusion in the review.

| Acronym | Study component | Inclusion Criteria | Exclusion criteria |
| --- | --- | --- | --- |
| P | Population | Migrant women aged 15 and over | - Population with a particular pathology<br>- Population exclusively or predominantly (i.e. >50%) male<br>- Age exclusively or predominantly (i.e. >50%) under 15 years |
| I | Intervention | Interventions aimed at empowerment in sexual health will be taken into consideration.<br>- Prevention, screening and treatment of sexually transmitted infections<br>- Screening for cervical and breast cancer<br>- Sex education<br>- Screening and care for sexual violence<br>- Contraception and birth control |  |
| C | Comparators | None defined |  |
| O | Outcomes | All outcomes concerning the effectiveness, acceptability and feasibility of sexual health empowerment interventions | Studies with economic criteria as the main outcome |
| S | Study design | - Randomised controlled trials and pilot randomised controlled trials<br>- Quasi-experimental studies<br>- Qualitative studies | - Any exclusively descriptive study<br>- Literature reviews, editorials, commentaries, books<br>- Observational studies |

PICOS: Population, intervention, comparator, outcome, and study design criteria (adapted from Satos et al., 2007)

**Additional file 1a: Search equations**

| Database | Search strategy | Filters |
| --- | --- | --- |
| **PubMed** | ((((((((((("sexual health"[Mesh] OR "reproductive health"[Mesh] OR "sexual health care" OR "reproductive health services"[Mesh] OR "sexual behavior"[Mesh] OR "gynecology"[Mesh] OR "prevention*" OR "sex offense*" OR "sexually transmitted disease*" OR "papanicolaou test »[Mesh] OR "Mammography"[Mesh] OR "mass screening"[Mesh] OR "contraception*" OR "early detection of cancer"[Mesh] OR "circumcision, female"[Mesh])<br>AND<br>((("Transients and Migrants"[Mesh] OR "emigrants and immigrants"[Mesh])<br>AND<br>((("intervention*" OR "health education"[Mesh])<br>AND<br>(((((("North America"[Mesh] OR "Europe"[Mesh]) OR "Oceania"[Mesh]) OR "China"[Mesh]) OR "Japan"[Mesh]) | Age (Adolescent: 13-18 years and older) |
| **Cochrane** | ("migrant"):ti,ab,kw OR ("immigrant"):ti,ab,kw OR<br>("refugee"):ti,ab,kw OR ("nomad"):ti,ab,kw OR<br>("transient"):ti,ab,kw<br>AND<br>("mutilation"):ti,ab,kw OR ("screening"):ti,ab,kw OR<br>("contraception"):ti,ab,kw OR ("sexual"):ti,ab,kw OR<br>("reproductive health education"):ti,ab,kw<br>AND<br>("North America"):ti,ab,kw OR ("Europe"):ti,ab,kw OR<br>("Oceania"):ti,ab,kw OR ("Japan"):ti,ab,kw OR<br>("China"):ti,ab,kw<br>AND<br>("intervention"):ti,ab,kw OR ("education"):ti,ab,kw | None |
| **Embase** | ('cancer prevention'/exp OR 'sexual crime'/exp OR<br>'female genital mutilation'/exp OR 'mammography'/exp<br>OR 'mass screening'/exp OR 'sexually transmitted<br>disease'/exp OR 'papanicolaou test'/exp OR<br>'contraception'/exp OR 'early cancer diagnosis'/exp OR<br>'contraceptive behavior'/exp OR 'gynecology'/exp OR<br>'sexual health'/exp)<br>AND<br>('migrant'/exp OR 'migration'/exp OR 'nomads'/exp OR<br>'refugee'/exp OR 'immigrant'/exp) | Age (Adolescent and older) |
|  | AND<br>('north america'/exp OR 'pacific islands'/exp OR 'china'/exp OR 'japan'/exp OR 'europe'/exp)<br>AND<br>('early intervention'/exp OR 'education'/exp OR 'patient care'/exp)<br>AND<br>([adolescent]/lim OR [adult]/lim OR [young adult]/lim OR [middle aged]/lim OR [aged]/lim OR [very elderly]/lim) |  |
| **Psychinfo/**<br>**Psycharticles** | ("sexual health" OR "reproductive health" OR "sexual health care" OR "reproductive health care" OR "sexual behavior" OR "gynecology" OR "sex offense*" OR "female genital mutilation*" OR "circumcision*" OR "sexually transmitted disease*" OR "papanicolou test" OR "mammograph*" OR "mass screening" OR "contraception" OR "contraceptive behavior" OR "early detection of cancer")<br>AND<br>("Transients and Migrants" OR "emigrants and immigrants" OR "refugee*" OR "nomad*") | Age (Adolescent and older) |
|  | AND<br>("intervention*" OR "education*")<br>AND<br>("North America" OR "Europe" OR "Oceania" OR "China" OR "Japan") |  |
| **Web of science** | ((((((((((((((ALL=("sexual health")) OR ALL=("reproductive health")) OR ALL=("sexual behavior")) OR ALL=("gynecology")) OR ALL=("gynaecology")) OR ALL=("sex offense*") OR ALL=("female genital mutilation")) OR ALL=("cancer prevention")) OR ALL=("cancer screening")) OR ALL=("sexually transmitted disease")) OR ALL=("sexually transmitted infection*") OR ALL=("papanicolou test")) OR ALL=("mammograph*") OR ALL=("contraception*") OR ALL=("circumcision female")) OR ALL=("early cancer diagnosis"))<br>AND.<br>((((ALL=(europe)) OR ALL=(oceania)) OR ALL=(north america)) OR ALL=(china)) OR ALL=(japan).<br>AND.<br>((((ALL=("migrant*")) OR ALL=("immigrant*")) OR | None |
|  | ALL=("nomad*")) OR ALL=("refugee*")) OR<br>ALL=("transient*")<br>AND.<br>((((ALL=("early intervention")) OR<br>ALL=("intervention*")) OR ALL=("education*")) OR<br>ALL=("health education") |  |
| **Cairn** | Santé sexuelle ET Migrant* ET Intervention | None |

**Additional file 1b : Grey literature and manual search**

| Database | Search strategy | Filters |
| --- | --- | --- |
| **ClinicalTrials.gov** | Combination of two groups of keywords:<br>- Migrant, refugee, nomad, transient<br>- « sexual health", "reproductive health",<br>"sexual health care", "reproductive health services",<br>"sexual behavior", "gynecology", "prevention*",<br>"sex offense*", "sexually transmitted disease*",<br>"papanicolaou test", "Mammography", "mass<br>screening", "contraception*", "early detection of | Sex : Female Locations :<br>"France" OR "China"<br>OR "Japan" OR<br>"United-States" OR<br>"Canada" OR<br>"Australia" OR "New-Zealand" OR<br>"Germany" OR "United<br>Kingdom" OR<br>"Denmark" OR |
|  | cancer", "circumcision, female". | "Finland" OR "Sweden"<br>OR "Greece" OR<br>"Spain" OR "Italia" OR<br>"Portugal" OR "Austria"<br>OR "Norway" |
| **SUDOC** | "santé sexuelle", "migrant*" | All words search |
| **Other websites:**<br>French National<br>Authority for<br>Health; WHO<br>; Institut<br>Convergence<br>Migration; French<br>College of<br>Gynaecology and<br>Obstetrics | Manual search | None |

